# Incidence and Pattern of Childhood Cancer in Addis Ababa, Ethiopia (2012 - 2017)

**DOI:** 10.1101/2023.04.04.23288123

**Authors:** Amanuel Belay, Ahmed Ali, Wondimu Ayele, Mathewos Assefa, Ahmedin Jemal, Eva J. Kantelhardt

## Abstract

**Background:** Cancer is becoming a major public health problem and a leading cause of death in children worldwide. However, little is known about the epidemiology of childhood cancer in Ethiopia. This study, therefore, assessed childhood cancer incidence patterns in Addis Ababa using the Addis Ababa city population-based cancer registry data from 2012-2017.

**Methods:** Invasive cancer cases diagnosed in ages 0-14 years from 2012-2017 were obtained from the Addis Ababa City population-based Cancer Registry. Cases were grouped according to the International Classification of Childhood Cancer, 3rd edition (ICCC-3) based on morphology and primary anatomic site. Age-standardized incidence rates (ASR) were calculated by the direct method using the world standard population.

**Results:** The overall average annual incidence rate during 2012-2017 in children was 84.6 cases per million, with rates higher in boys (98.97 per million) than in girls (69.7 per million). By age, incidence rates per million increased from 70.8 cases in ages 0-4 years to 88.4 cases in ages 5-9 years to 110.0 cases 10-14 years. Leukemia was the most common childhood cancer in both boys (29.1%) and girls (26.8%), followed by lymphoma in boys (24.7%) and renal tumours (13.1%) in girls. The overall cancer incidence rate decreased from 87.02 per million in 2012 to 51.07 per million in 2017.

**Conclusion:** The burden of childhood cancer is considerably high in Addis Ababa, and its profile is generally similar to those from other parts of sub-Saharan Africa except that renal tumour is the 2^nd^ most commonly diagnosed cancer in girls in Addis Ababa. Future studies should examine risk factors for the occurrence of childhood cancer in Addis Ababa and other parts of Ethiopia.

## Introduction

Childhood and adolescent cancers contribute significantly to the global cancer burden [1]. Childhood cancer incidence has increased by 13% globally [2]. Each year more than 300000 Children and adolescents are diagnosed with cancer worldwide.[3]. Of children who develop cancer, approximately 90% occur in low- and middle-income countries More than 80% of diagnosed cases of childhood cancer occur in low-income and middle-income countries[4]. Due to limited access to diagnostics and treatment [5] the survival rates are less than 30% in lower-middle-income and low-income countries compared to over 80% in high-income countries (HIC)[6].

The Addis Ababa cancer registry, established in 2011, is the only population-based cancer registry in the country, and childhood cancer cases account for about 4.6% of the total cases [7]. According to clinical record findings from Tikur Anbessa Specialized Hospital, which hosts the only oncology facility in the country[8], most childhood cancer cases present at a late stage of the disease when the chance of survival is poor [9].

Few studies reported on childhood cancer patterns in Ethiopia. Further, all these studies were based on a medical chart review from a single institute rather than a population-based cancer registry[10-12]. Ethiopia has adopted a comprehensive National Action Plan for the Prevention and Control of Chronic Non-Communicable Diseases, and the expansion of cancer treatment services is underway[9]. And epidemiological data are important for estimating and proper allocation of resource needs and health policy prioritization [1].

Therefore, this research has used population-based cancer registry data from Addis Ababa to assess the incidence rate and pattern of childhood cancer in Addis Ababa city. Findings from this research can be used to plan cancer control actions, health care, and allocation of resources for developing effective programs aiming at the control, prevention, diagnosis, and treatment guidelines of childhood cancer in Ethiopia.

## Methods

### Study design and data sources

In this population-based registry study, Cancer incidence data from 2012-2017 were obtained from Addis Ababa City Cancer Registry. The Addis Ababa City Cancer Registry is the first population-based Cancer Registry (AAPBCR) in the country and it was established in September 2011 at Radiotherapy Center, Tikur Anbessa Specialized Hospital, School of Medicine and Addis Ababa University. The registry actively collects all new cancer patients who are residents of Addis Ababa from 20 collaborating institutions (pathology, oncology and radiotherapy facilities). The population at risk was obtained from the Ethiopian central statistical agency.

### Statistical analysis

The diagnosis was classified according to the 3rd edition of the International Classification of Diseases for Oncology (ICCC-3)[13]. Age-specific incidence rates (ASR) were calculated for three 5-year age groups (0–4 years, 5–9 years, and 10–14 years). Incidence rates were calculated as the average annual number of cases per million child years. Age-standardized rates (ASR) were calculated by the direct method using the world standard population[14]. Incidence of sex ratios was calculated by dividing the incidence among males by that in a female.

Statistical analyses were done using Stata/SE (version 14) [15].

## Results

During the period 2012-2017, 395 childhood cancers were diagnosed in Addis Ababa, Ethiopia. Two hundred twenty-seven (57.5%) of the patients were males and 168 (42.5%) of the patients were females. The median age was 6 (IQR, 3-11). A majority (39.2%) of the patients were between the age group of 0-4 years, followed by 10-14 (31.1%) years age group and 5-9 (29.6%) years age group respectively.

Leukemias were the most common type of cancer diagnosed among Addis Ababa children, with an incidence of 111 (28.1%). The second most common childhood cancers were Lymphomas, with a frequency of 75(19%), followed by Soft tissue and other extraosseous sarcomas with 51(12.9%), Renal Tumors with 39 (9.9%) and other malignant epithelial neoplasms and malignant melanomas with 28 (7.1%).

Childhood cancer types were found different for males and females (Table 1) (Stable 1). In males, Leukemias (29.1%) were the most common cancer followed by Lymphomas (24.7%), Soft tissue and other extraosseous sarcomas (13.7%), Malignant bone tumours (7.5%) and Other malignant epithelial neoplasms and malignant melanomas (6.2%) respectively. In contrast, in females, leukaemias (26.8%) were the most common cancer followed by Renal tumours (13.10%), Soft tissue and other extraosseous sarcomas (11.9%), Lymphomas (11.3%) and Other malignant epithelial neoplasms and malignant melanomas (8.3%).

**Table 1:** Number of childhood cancer in Addis Ababa from 2012-2017 by diagnostic group of the International Classification of Childhood Cancer-third edition and sex.

| Types of Malignancy | Male | Female | Total |
| --- | --- | --- | --- |
|  | N(%) | N(%) | N(%) |
| **All cases** | 227(100.0) | 168(100.0) | 395(100.0) |
| **Leukemias, myeloproliferative diseases, and myelodysplastic diseases** | 66(29.0) | 45(26.8) | 111(28.1) |
| **Lymphomas and reticuloendothelial neoplasms** | 56(24.7) | 19(11.3) | 75(19) |
| **Soft tissue and other extraosseous sarcomas** | 31(13.7) | 20(11.9) | 51(12.9) |
| **Renal tumours** | 12(5.3) | 22(13.1) | 39(9.9) |
| **Other malignant epithelial neoplasms and malignant melanomas** | 14(6.2) | 14(8.3) | 28(7.1) |
| **Malignant bone tumours** | 17(7.5) | 9(5.4) | 26(6.6) |
| **Retinoblastoma** | 7(3.1) | 12(7.1) | 19(4.8) |
| **CNS and miscellaneous intracranial and intraspinal neoplasms** | 7(3.1) | 6(3.6) | 13(3.3) |
| **Germ cell tumours, trophoblastic tumours, and neoplasms of gonads** | 1(0.4) | 11(6.5) | 12(3.0) |
| **Neuroblastoma and other peripheral nervous cell tumours** | 3(1.3) | 7(4.2) | 10(2.5) |
| **Other and unspecified malignant neoplasms** | 5(2.2) | 2(1.2) | 7(1.8) |
| **Hepatic tumours** | 3(1.3) | 1(0.6) | 4(1.0) |

The range of tumour types varied markedly with age groups (Table 2). In the age group 0-4, Leukemias were the most common cancer representing 26% of all cases. However the proportion was higher in the age group 5-9 representing 32.5% of cases but, in the age group 10-14, the proportion of Leukemias was 22.8% of all cases. Renal tumours were the second most frequent tumours in the age group 0-4 representing 16.8% of cases however the proportion of Renal tumour cases was 7.7% and 3.3% in the age group 5-9 and 10-14 respectively. Soft tissue sarcomas were the third most common cancer in the age group 0-4 (13.5%) and 5-9 (11.1%). Bone tumours represented 0.6% of all cases in the age group but were the third most common cancer in the age group 10-14.

**Table 2:** Number of childhood cancer in Addis Ababa from 2012-2017 by diagnostic group of the International Classification of Childhood Cancer-third edition and age group.

| Diagnosis group | Age group |  |  |
| --- | --- | --- | --- |
|  | 0-4 | 05-09 | 10-14 |
|  | N (%) | N (%) | N (%) |
| **All cases** | 155 (39.2) | 117 (29.6) | 123 (31.1) |
| **Leukemias, myeloproliferative diseases, and myelodysplastic diseases** | 45 (11.4) | 38 (9.6) | 28 (7.1) |
| **Renal tumours** | 26 (6.6) | 9 (2.3) | 4 (1.0) |
| **Soft tissue and other extraosseous sarcomas** | 21 (5.3) | 13 (3.3) | 17 (4.3) |
| **Lymphomas and reticuloendothelial neoplasms** | 18 (4.6) | 29 (7.3) | 28 (7.1) |
| **Retinoblastoma** | 18 (4.6) | 1 (0.3) | 0 |
| **Other malignant epithelial neoplasms and malignant melanomas** | 10 (2.5) | 4 (1.0) | 14 (3.5) |
| **Neuroblastoma and other peripheral nervous cell tumours** | 4 (1.0) | 5 (1.3) | 1 (0.3) |
| **Germ cell tumours, trophoblastic tumours, and neoplasms of gonads** | 4 (1.0) | 1 (0.3) | 7 (1.8) |
| **Other and unspecified malignant neoplasms** | 4 (1.0) | 2 (0.5) | 1 (0.3) |
| **CNS and miscellaneous intracranial and intraspinal neoplasms** | 3 (0.8) | 5 (1.3) | 5 (1.3) |
| **Hepatic tumours** | 1 (0.3) | 3 (0.8) | 0 |
| **Malignant bone tumours** | 1 (0.3) | 7 (1.8) | 18 (4.6) |

The overall incidence rate of childhood cancer from 2012-2017 was 84.6 cases per million with an average of 65.8 cases per year. The incidence rate in boys was 98.97 cases per million and that of girls was 69.7 cases per million. The sex ratio for all childhood cancer incidence was 1.42. The incidence rate was higher among children aged 10-14 with 100 cases per million followed by children aged 5-9 and 0-4 with 79 cases per million and 76.6 cases per million respectively.

The overall cancer incidence rate decreased from 85.4 per million in 2012 to 50.4 per million in 2017 (Table 3). The incidence rates were higher among boys than girls from 2012 to 2017. Age group 10-14 had the highest incidence rate in both 2012 and 2017. However, the incidence rate was higher among the age group 5-9 in the year 2013 (124.3 per million). Leukemia’s had the highest incidence rate in both the years 2012 and 2017 (Table 4). Meanwhile, in the year in the year 2015, Lymphoma’s had the highest incidence rate with 17.5 per million.

**Table 3:** Childhood cancer incidence by sex and age group from 2012 – 2017.

|  | 2012 |  | 2013 |  | 2014 |  | 2015 |  | 2016 |  | 2017 |  |
| --- | --- | --- | --- | --- | --- | --- | --- | --- | --- | --- | --- | --- |
|  | N | ASR | N | ASR | N | ASR | N | ASR | N | ASR | N | ASR |
| **Over All** | 62 | 85.4 | 80 | 106.4 | 86 | 108.8 | 63 | 76.8 | 60 | 71.7 | 44 | 50.4 |
| **Male** | 36 | 102.8 | 49 | 132.7 | 45 | 116.2 | 35 | 86 | 30 | 72.8 | 32 | 73.6 |
| **Female** | 26 | 68.3 | 31 | 80.3 | 41 | 102.4 | 28 | 67.7 | 30 | 70.8 | 12 | 27.5 |
| **Age Group** |  |  |  |  |  |  |  |  |  |  |  |  |
| **00-04** | 22 | 68.8 | 31 | 93.9 | 37 | 108.5 | 27 | 76.6 | 20 | 54.9 | 18 | 47.8 |
| **05-09** | 17 | 85.9 | 27 | 124.3 | 23 | 96.4 | 18 | 68.8 | 20 | 69.6 | 12 | 37.6 |
| **10-14** | 23 | 106.9 | 22 | 103.2 | 26 | 123.0 | 18 | 85.9 | 20 | 96.3 | 14 | 68.0 |

<sup>a</sup>ASR = Age Standardized Incidence Rate

**Table 4:**
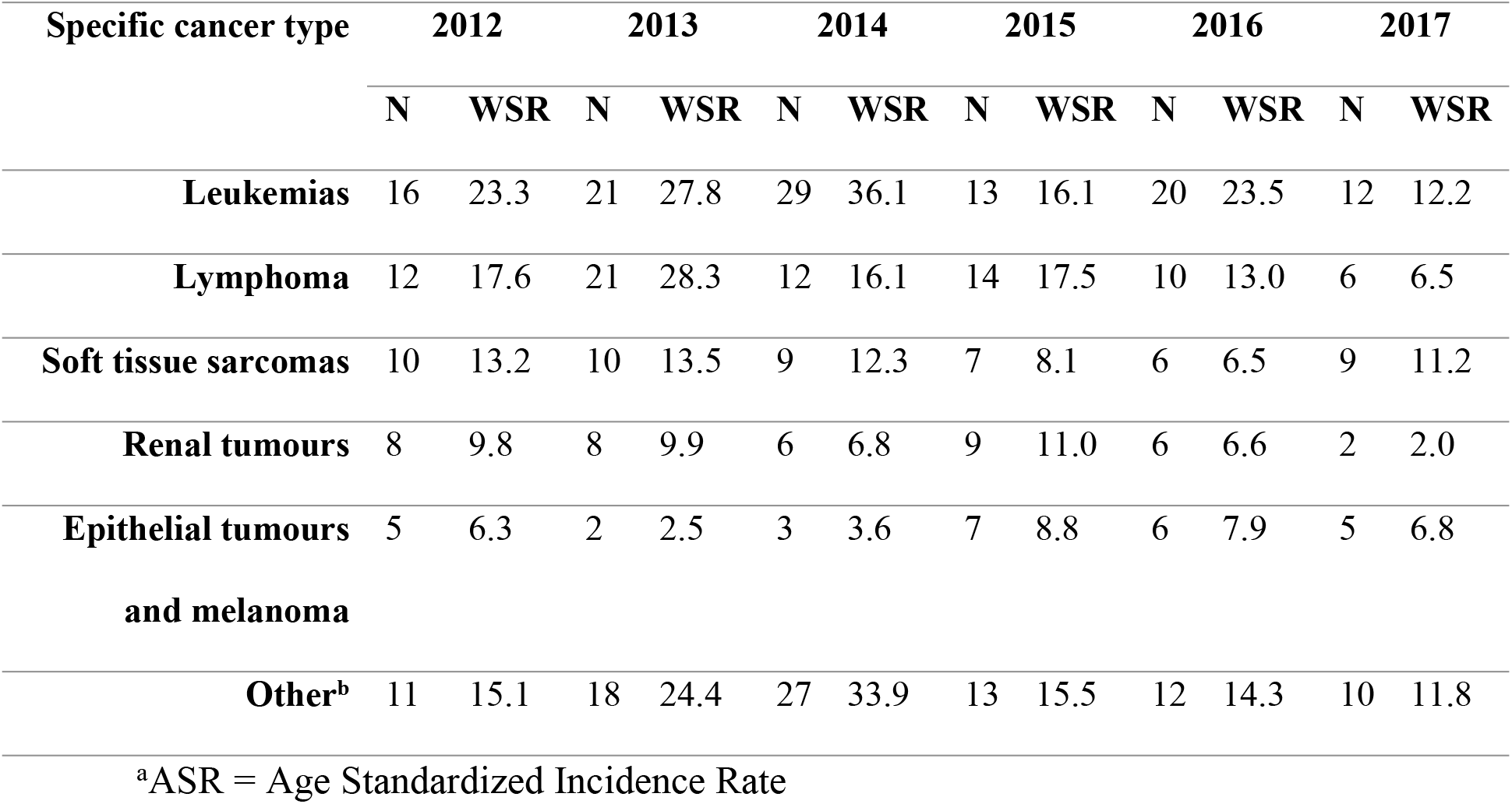
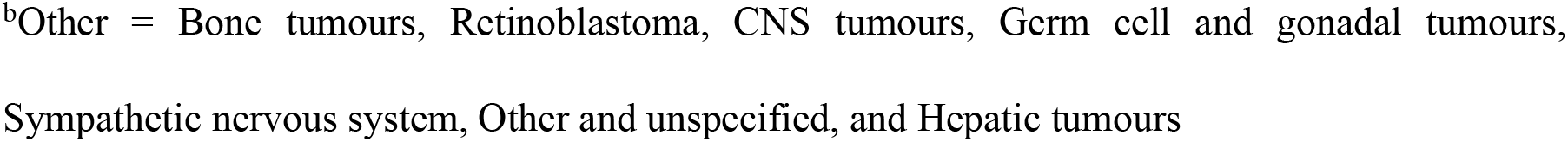
Childhood cancers incidence by diagnostic group of the International Classification of Childhood Cancer-third edition.

## Discussion

This is the first population-based childhood cancer study done in Ethiopia. The key findings from this study showed that the overall incidence rate of childhood cancer from 2012-2017 was 81.9 cases per million.

In Addis Ababa, the most common cancer was leukaemias followed by Lymphoma and Soft tissue and other extraosseous sarcomas respectively. In contrast to our finding at Tikur Anbessa specialized Hospital the most common malignancies were Wilm’s tumour (24.7%) followed by leukaemia (19.5%) and lymphomas (14.3%) respectively from January 2005 to December 2006 [12]. And at Gondar University Hospital, 60.6% of the reported childhood cancers were haematological malignancies, followed by Wilms tumour [11]. Another study in Gonder hospital showed that the most common malignancies in children to be Lymphoma, Wilma’s tumour, and Retinoblastoma respectively[10]. The observed difference in the incidence of leukaemia might be due to, the difficulty in diagnosing. leukaemia presents with protean signs and symptoms that resemble those of infection, in which early death can occur before cancer is suspected or diagnosed [16].

The observed distribution was different compared to other African centuries. In Africa, the most common childhood cancer is Lymphomas, [17] nephroblastoma, Kaposi sarcoma, and retinoblastoma[17]. most malignancies in Africa are related to infections like Malaria, EBV, HIV/AIDS, and human herpesvirus 8 (HHV8) [18]. Similar to our findings, in Tunisia, Leukaemias, and Lymphomas are reported as the most common childhood cancers respectively [19]. However, in Tunisia, CNS is the third most common cancer in children. In Blantyre, Malawi from 2008-2010, the most common childhood cancer was Burkitt lymphoma[20]. This was followed by Wilms’ tumour and Kaposi sarcoma. In Namibia Leukaemias and retinoblastomas were the most common tumours, with renal tumours, soft tissue sarcomas and lymphomas following in frequency[21]. Due to a lack of adequate diagnostic and treatment facilities leukaemias and brain cancers are underdiagnosed [22]. However, other childhood cancers, including infection-related cancers such as Kaposi sarcoma, Burkitt lymphoma, Hodgkin lymphoma, and hepatocellular carcinoma, and also two common embryonal cancers retinoblastoma and nephroblastoma incidence rates in Africa are higher than those in high-income countries [22].

Brain tumours are the second most common malignancy reported in children worldwide [2] and in some countries [23-25] or the third most common malignancy [26-28]. In contrast, In Ethiopia brain tumours represents only 3% of identified childhood cancers [29]. In Addis Ababa, CNS cancer is the eighth most common cancer in Addis Ababa. The differences may be due to environmental or genetic differences[30]. In Addition, diagnosing brain tumours in Africa is difficult due to the limited availability of necessary special diagnostic tools, financial implications and in most cases the wrong assumption of differential diagnoses such as cerebral malaria, tuberculous meningitis, bacterial meningitis, and others [17].

In Addis Ababa, the overall age-standardized incidence rates of childhood cancer were 81.9 cases per million. The incidence rate in boys was 95.5 per million and that of girls was 68.7 per million. Globally, the overall childhood cancer incidence rate was 140.6 per million person-years in children aged 0–14 years [2]. In South Africa, the overall age-standardized incidence rate (ASR) for 2000–2006 was 45.7 per million children [31]. In Namibia, the overall childhood cancer incidence rate for 2003-2010 was 29.4 per million [21]. The observed childhood cancer incidence in this study was higher than the incidence observed in South Africa (45.7 per million)[31] and Namibia (29.4 per million)[21]. However, it was lower when compared to the global childhood cancer incidence rate (140.6 per million) and other countries like Zimbabwe (130.0 per million) [32], Uganda (147.2 per million) [32], Spain (155.8 per million) [24], Thailand (74.9 per million) [27], and Korea (134.9 per million) [33]. This difference in the incidence rate may be attributable to diversity registration practices, and actual racial and geographical differences. Such differences may be the result of genetic tendency, early or delayed exposure to infectious agents, and other environmental factors. Some studies have stated that the incidence of childhood malignancies in low-income countries is lower than in industrialized countries[16]. Registration of childhood cancer requires the identification of symptoms, rapid access to a pediatric cancer unit, a correct diagnosis, and a data management foundation. In low-income countries, where these services are lacking, some children with cancer may die before diagnosis and registration [16]

The incidence of childhood cancer is not expected to increase in Africa. However, an increase in the number of reported cases might occur with the improvement of the quality of registries and the increase in awareness, knowledge, diagnostic tools, and affordability[17]. With the decrease in Kaposi sarcoma, cancer-correlated HIV, and an increase in leukaemia cancer correlated with a higher level of development a change in the trend of childhood cancer is expected in Africa [17, 34]. In this study, a decreasing trend in the incidence of childhood cancer was observed. In contrast to our finding a study done at Tikur Anbessa spatialized Hospital and Gondar University Hospital showed an increased incidence of childhood cancer [11, 12]. The observed decreasing incidence rate in this study might be due to, patients being diagnosed at a health facility that is not the source of the cancer registry, misdiagnosis, lack of quality and affordable diagnostic tools, or lack of quality in the cancer registry.

## Conclusion

The overall incidence rate of childhood cancer from 2012-2017 was 81.9 cases per million. The incidence rate showed a decreasing pattern in Addis Ababa, both in males and females. Although we have a limited understanding of what is driving these changes, the incidence pattern seen in this study may reflect random variation or changes in reporting or diagnosis. Accurate Childhood cancer incidence data are needed to drive continuous improvements in the quality of care and for policymakers to inform priority setting and planning decisions. This research highlights the need for more research into the epidemiology and aetiology of childhood cancer to implement adequate cancer control and prevention programs.

## Data Availability

The data that support the findings of this study are available from Addis Ababa Cancer Registry but restrictions apply to the availability of these data, which were used under license for the current study, and so are not publicly available. Data are however available from the authors upon reasonable request and with permission of the Addis Ababa Cancer Registry.

## Acknowledgements

We acknowledge the staff of Addis Ababa Cancer for their cooperation during data collection. We would also like to acknowledge, the American Cancer Society and Martin Luther University of Halle, Germany for supporting the registry financially.

## Supporting Information

**S1 Table: Number of childhood cancer in Addis Ababa from 2012-2017 by diagnostic group of the International Classification of Childhood Cancer-third edition and sex**

**S2 Table: Number of childhood cancer in Addis Ababa from 2012-2017 by diagnostic group of the International Classification of Childhood Cancer-third edition and age group**

